# Phenol-chloroform-based RNA purification for detection of SARS-CoV-2 by RT-qPCR: comparison with automated systems

**DOI:** 10.1101/2020.05.26.20099440

**Authors:** Henrik Dimke, Sanne L. Larsen, Marianne N. Skov, Hanne Larsen, Gitte N. Hartmeyer, Jesper B. Moeller

## Abstract

The outbreak of severe acute respiratory syndrome coronavirus 2 (SARS-CoV-2) has rapidly reached pandemic levels. Sufficient testing for SARS-CoV-2 remains essential for tracking and containing the rapid spread of the virus. However, due to increased global demand, kits and proprietary reagents for RNA extraction are limited, which markedly reduce SARS-CoV-2 testing capabilities in many countries. Here, we explore the use of conventional acid guanidinium thiocyanate-phenol-chloroform (AGPC)-based RNA purification as an alternative to commercial automated systems for detection of SARS-CoV-2 by RT-qPCR. 87 clinical oropharyngeal or nasopharyngeal swab specimens were extracted by AGPC and compared to the commercial platforms, the Promega Maxwell^®^ RSC 48 instrument for automated RNA extraction and the fully integrated diagnostic system, the Cobas^®^6800 apparatus. Our results show that RNA extracted using the AGPC method is fully comparable to modern automated systems regarding analytical sensitivity, specificity and accuracy with respect to detection of SARS-CoV-2 as evaluated by RT-qPCR. Moreover, we find that the AGPC method is easily scalable and implemented in conventional laboratories. Taken together, these data identify conventional AGPC-based RNA extraction as a low cost and suitable alternative to automated systems for the detection of SARS-CoV-2, when automated systems, kits and reagents are not readily available.

## Introduction

The severe acute respiratory syndrome coronavirus 2 (SARS-CoV-2) causing the coronavirus disease 2019 (COVID-19) has rapidly reached pandemic levels, with COVID-19 related morbidities and mortalities rising in many countries (1-3). All efforts are needed to regain control and one important aspect is tracking the spread of infections, both in the healthcare system and in the general public. However, the increased spread of the virus has markedly increased global demand for materials and reagents needed for adequate testing. The limited supply of commercial RNA extraction kits, consumables and reagents has posed serious limitations on testing capacities in many countries, especially those relying on automated systems where commercial kits are indispensable. Furthermore, commercial systems and kits are expensive and not readily available in all countries. Without the ability to test more widely it is difficult to adequately evaluate the spread of the disease. In the wake of this, several new methods have been proposed to overcome the bottleneck posed by RNA purification in an effort to detect the novel coronavirus (SARS-CoV-2) and clinically diagnose the COVID-19 (4, 5). As such the acid guanidinium thiocyanate-phenol-chloroform (AGPC) method of RNA extraction has recently been found suitable to allow for SARS-CoV-2 PCR detection (5). However, it is unclear how it compares to automated systems currently running at clinical laboratories and if levels of analytical sensitivity, specificity and accuracy are comparable. A good accuracy is especially important since false negative samples could lead to inadvertent spread of the COVID-19 disease within the healthcare systems and general communities.

The AGPC is a simple method used in many laboratories worldwide. The method itself has a long-known track record (6) and relies on an acid guanidinium thiocyanate-phenol mixture that can be used to extract RNA with the addition of chloroform. Commercially mixed reagents needed for this step are readily available from several vendors as e.g. TRIzol™ (Invitrogen) and TRI Reagent^®^ (Sigma-Aldrich, now Merck) or the reagents can be produced locally from base chemicals at a low cost (6). The method is simple to implement in conventional laboratory settings and is already routinely used at many research institutions. One caveat in comparison with automated systems is the additional hands-on sample preparation time needed. For COVID-19 testing it is currently unknown whether the AGPC-based RNA extraction method can perform to a level similar to automated testing systems, especially when reagents for the latter systems are sparse.

We therefore aimed to compare the AGPC method to automated systems commonly used in clinical detection of SARS-CoV-2 and to evaluate its suitability as a replacement for conventional automated systems when shortages of proprietary materials are experienced or not readily available. Furthermore, we aimed to explore whether the AGPC extraction method could be used for detection of SARS-CoV-2 on a larger scale. Our current data shows that accuracy of this method is fully comparable to automated systems. This is important since the AGPC is a simple method used in laboratories worldwide to extract RNA and commercially formulated reagents needed for this technique are readily available or can be easily prepared from basic laboratory chemicals at a low cost.

## Materials and methods

### Sample material

Sample material was obtained from oropharyngeal or nasopharyngeal swabs collected in ESwab™ tubes containing 1 ml of liquid Amies medium (Copan). Specimens were obtained from patient material undergoing routine diagnostic analyses at the Department of Clinical Microbiology, Odense University Hospital. The samples used for this study were chosen based on SARS-CoV-2 status from the routine diagnostic analyses and to reflect a broad range of viral titers in the sample material as determined by the *SARS-CoV-2 E* gene (Ct value range 16-38) using the Cobas^®^6800 system (Roche). To distinguish between true negative results and reactions affected by inadequate RNA isolation, presence of PCR inhibitors or instrument failure, Nobilis ND C2 vaccine (NDV) against Newcastle Disease (Nobilis) was added to the ESwab™ media as an internal control prior to RNA purification using the AGPC method and the Maxwell^®^ RSC 48 instrument (Promega). The amount of added NDV had previously been titrated to yield a Ct value ~27.5 as measured by RT-qPCR and routinely used for evaluating sample quality after RNA isolation at the Department of Clinical Microbiology. To determine a Ct cutoff value for NDV after purification with the Maxwell^®^ RSC 48 and the AGPC methods, 24 clinical swab samples with known SARS CoV-2 status based on the Cobas^®^6800 system were processed using both methods and the data for the inhouse NDV and SARS-CoV-2 RT-qPCR assays were evaluated. Based on these data and applying a precautionary principle the NDV Ct cutoff value was determined to be <29.5.

### Cobas^®^6800 *system*

The fully automated IVD-CE-labelled Cobas^®^6800 system (Roche Diagnostics, Basel, Switzerland) was used in this study (set as gold standard) for the evaluation of Maxwell^®^ and AGPC methods of RNA purification. Testing was performed using the Cobas^®^ SARS-CoV-2 test assay with proprietary primers directed against the *SARS-CoV-2 ORF1* and E-gene. 400 μl ESwab™ sample media was added to the Cobas^®^ machine and eluted in a final volume of 50 μl. From this, 27 μl of eluted sample was added to 25 μl of Cobas^®^ SARS-CoV-2 PCR mix.

### Maxwell^®^ RSC 48 automated RNA extraction

RNA extraction was performed with the Maxwell^®^ RSC Viral Total Nucleic Acid Purification Kit (Promega) using the Maxwell^®^ RSC 48 Instrument according to the manufacturer’s recommendation, without the initial heat incubation step. In brief, lysis buffer, Proteinase K and internal controls were added to the cartridge and 200 μl of ESwab™ sample media was added. The extracted RNA was eluted in a total volume of 50 μl.

### The acid guanidinium thiocyanate-phenol-chloroform extraction method

RNA extraction was carried out according to the instructions of the Tri Reagent^®^ manufacturer with minor modifications. In brief, 200 μl of the sample specimens were aliquoted into sterile 1.5 ml test tubes containing 800 μl of TRI Reagent^®^ (Catalog No. T9424, Sigma-Aldrich). This step was performed under Class II conditions, while the remainder of the RNA extraction, after inactivation of the virus, was performed in a conventional laboratory. Once mixed and transported to the conventional laboratory, the test tubes with sample material and Tri Reagent^®^ were added 200 μl chloroform and mixed by vortexing (5 sec. at max speed). Samples were then incubated for 2 minutes at room temperature and subsequently centrifuged at 14,000g for 15 minutes (4°C). Only the aqueous phase (500 μl) of the resulting mixture located at the top of the tube was processed further, while the remainder of the Tri Reagent^®^/chlorofom mixture was discarded. The aqueous phase was pipetted into a new tube containing 2 μl of GlycoBlue^TM^ (Catalog No. AM9515 ThermoFisher). 600 μl of isopropanol was then added to the tubes containing the aqueous phase, after which the samples were mixed by vortexing (3 sec at max speed) and then incubated at room temperature (20-25°C) for 20 minutes. The samples were then centrifuged at 14,000g for 15 minutes (4°C), the supernatant removed while making sure the blue RNA pellet remained at the bottom of the tube. The RNA pellets were then washed in 1 ml of 75% Ethanol, vortexed (3 sec. at max speed) and centrifuged at 8,000g for 5 minutes (4°C). The supernatants were then removed while making sure that the RNA pellets remained at the bottom of the tubes. After removal of the Ethanol, samples were incubated at room temperature (20-25°C) for 10 minutes with the lid open to allow excess ethanol to evaporate. Subsequently the RNA pellets were resuspended in 30 μl of RNAse-free water and heated to 60°C for 5 minutes on a heating block. RNA samples were then vortexed 3 sec. and then centrifuged briefly (10 sec. on table centrifuge) to collect all the liquid at the bottom of the tube. Samples were stored at 4°C before transported on ice to the Department of Clinical Microbiology for further analysis.

### SARS-CoV-2 RT-qPCR analysis

The SARS-CoV-2 was detected according to the real-time PCR protocol established by Corman et al. (7). Detection of the internal control Newcastle disease virus NDV was performed using primers and probe sequences kindly provided by dr. Kurt J. Handberg, Department of Clinical Microbiology, Skejby, Denmark. In brief, TaqMan™ Fast Virus 1-Step Master Mix (ThermoFisher) was used for the amplification reaction. A final concentration 1000 nmol of primers and 200 nmol of probes were added to a total reaction volume of 20 ul, containing 6 μl of RNA (purified RNA obtained from either Maxwell^®^ automated RNA extraction or from AGPC extraction). Samples were analyzed on a LightCycler®480 II (Roche) using the following program 50°C for 5min, 95°C for 20sec followed by 45 cycles of 95°C for 15sec and 60°C for 1min.

E_Sarbeco_F1: ACAGGTACGTTAATAGTTAATAGCGT

E_Sarbeco_R2: ATATTGCAGCAGTACGCACACA

E_Sarbeco_P1: FAM_ACACTAGCCATCCTTACTGCGCTTC G_BHQ1

NDV-F: 5’-CAC TGT CGG CAT TAT CGA TGA-3’

NDV-R: 5’-GAG CAT CGC AGC GGA AA-3’

NDV-Probe: 5’-FAM-CCC AAG CGC GAG TTA-MGB-3’

### Comparison tests and Statistics

A total of 87 clinical sample specimens were chosen based on SARS-CoV-2 status from the Cobas^®^6800 system and used to evaluate the analytical sensitivity, specificity and accuracy of our in-house SARS-CoV-2 RT-qPCR assay after RNA purification using the Maxwell^®^ RSC 48 and AGPC methods. Only samples yielding Ct values below 29.5 for the internal control NDV assessed by RT-qPCR on the Maxwell^®^ and TRI Reagent^®^ platforms were included in the analyses. Results obtained on the Cobas^®^6800 system for the SARS-CoV-2 *E* gene were compared to the results obtained using our in-house RT-qPCR assay for the SARS-CoV-2 *E* gene. 95% confidence intervals and Pearson correlation coefficients (r) were calculated using Prism 8.3 (Graphpad Software).

## Results

### The AGPC method delivers high analytical sensitivity, specificity and accuracy for SARS-CoV-2 testing

To evaluate whether conventional AGPC based extraction of RNA could serve as a viable alternative to automated systems with respect to reliability and accuracy, we isolated RNA using the AGPC method from 87 clinical specimens (oropharyngeal or nasopharyngeal swabs) with known SARS-CoV-2 status (57 positive and 30 negative), and performed a side-by-side comparison with the identical samples extracted on a Maxwell^®^ RSC 48 instrument. The samples used had previously been analyzed using the state-of-the-art fully integrated Cobas^®^6800 diagnostic system capable of analyzing patient specimens from sample to test result without manual interference, using proprietary primers directed against the *SARS-CoV-2 ORF1* and *E*-gene. Analysis of the sample specimens with RNA extracted by the Maxwell^®^ RSC 48 instrument showed a 98.2% sensitivity, 96.4% specificity and 97.6% accuracy, but also reported a single false positive and a false negative sample compared to the Cobas^®^6800 system (Figure 1A). Importantly, analysis of the sample specimens using the AGPC method for RNA extraction displayed a 98.0% sensitivity, 100% specificity and 98.8% accuracy, with no false positive and only 1 false negative compared to Cobas^®^6800 system.

**Figure 1:**
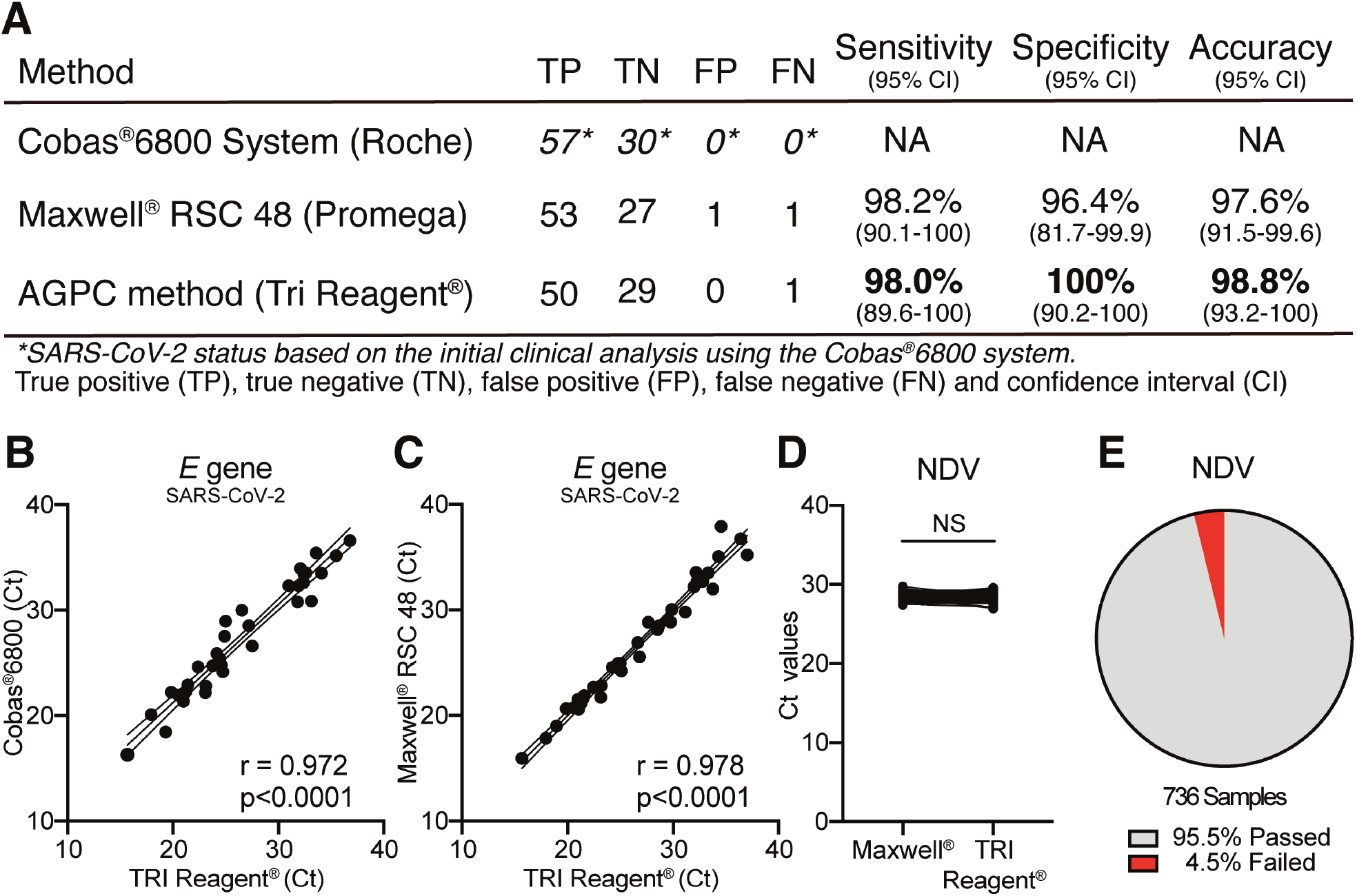
Phenol-chloroform extraction of RNA is a viable alternative to automated systems, showing similar levels of sensitivity, specificity and accuracy. **(A)** 87 patient specimens with known SARS-CoV-2 status based upon routine testing for SARS-CoV-2 using the Cobas^®^6800 platform were compared to results reported for the inhouse SARS-CoV-2 RT-qPCR assay after RNA isolation using the Maxwell^®^ RSC 48 instrument or TRI Reagent^®^. Only RNA specimens passing the internal control for the Newcastle Disease Virus vaccine strain (NDV, Ct values <29.5) were used for the analyses (Maxwell^®^; 82 samples, TRI Reagent^®^; 80 samples). True positive (TP), true negative (TN), false positive (FP), false negative (FN), confidence interval (CI). Sensitivity is defined as the probability a test result is positive for a SARS-CoV-2 positive sample. Specificity is defined as the probability a test result is negative for a SARS-CoV-2 negative sample. Accuracy is defined as the probability a patient sample is correctly evaluated for SARS-CoV-2. **(B)** Diagram showing a highly significant correlation (r = 0.970, p<0.001) between obtained Ct values for the SARS-CoV-2 *E* gene in SARS-CoV-2 positive specimens when assessed by RT-qPCR using the Cobas^®^6800 platform and the in-house SARS-CoV-2 RT-qPCR assay after RNA isolation using TRI Reagent^®^. **(C)** Diagram showing a highly significant correlation (r = 0.978, p<0.001) between obtained Ct values for the SARS-CoV-2 *E* gene in SARS-CoV-2 positive specimens when assessed by RT-qPCR using the in-house RT-qPCR assay after RNA isolation using the Maxwell^®^ RSC 48 instrument and TRI Reagent^®^. **(D)** Side-by-side comparison of Ct values obtained for the internal control NDV when assessed by RT-qPCR using identical patient specimens from which RNA was isolated using the Maxwell^®^ RSC 48 instrument or the AGPC method. Not significant (NS). **(E)** Pie chart showing the average rate of sample specimens that pass the NDV internal control for RNA extraction and sample quality after isolation of the RNA using the AGPC pipeline (n = 736).

### AGPC based RNA extraction allows for SARS-CoV-2 E-gene detection comparable to automated systems

To further validate our findings, we performed a direct comparison of the Ct values measured for the *SARS-CoV-2 E* gene in the SARS-CoV-2 positive sample specimens after AGPC extraction of the RNA, to the Ct values from identical samples reported using the Cobas^®^6800 system and the Maxwell^®^ RSC 48 instrument (Figure 1B and C), respectively. The Ct values attained for the *SARS-CoV-2 E* gene using the AGPC method showed a highly significant correlation to those reported for the Cobas^®^6800 system (r=0.97, p<0.0001) and the Maxwell^®^ RSC 48 instrument (r=0.98, p<0.0001).

To distinguish between true negative results, and reactions affected by inadequate RNA isolation, presence of PCR inhibitors or instrument failure, all sample specimens used for RNA extraction via the AGPC method and the Maxwell^®^ RSC 48 instrument were added Nobilis ND C2 vaccine (NDV) prior to RNA isolation. Thus, to assess the purity, yield and efficiency of the RNA extraction using the AGPC method we compared the Ct values reported for the NDV to those attained for the same identical samples extracted using the Maxwell^®^ RSC instrument. Direct comparison of the Ct values revealed no significant differences in Ct values for NDV between the AGPC method and the Maxwell^®^ RSC 48 instrument (Figure 1D).

### The AGPC method is easily scalable and allows for reliable RNA extraction in a conventional laboratory setting

The Department of Molecular Medicine at University of Southern Denmark is currently aiding the Department of Clinical Microbiology in detection of SARS-CoV-2 using the AGPC method. To examine the reliability of the AGPC-based isolation method in large scale, we set up a pipeline for RNA isolation. Using standard laboratory equipment set up in an empty teaching laboratory, we have adapted and modified the standard AGPC protocol for use in a large-scale RNA extraction pipeline (**Supplemental Figure 1 – Pipeline setup**). Taking an iterative approach, we have scaled up the number of samples processed daily, which has reduced the number of samples specimens that fail the NDV control to 4.5% (Figure 1E). The full protocol for this setup has been described in detail in supplemental materials (**Supplemental data – Detailed protocol**) and large-scale extraction can easiest be run with 5 technicians with the potential output of 400-600 samples specimens processed per day (7.5 hrs./day).

## Discussion

We find that the AGPC method is a reliable method for RNA extraction fully comparable to automated systems with respect to detection of SARS-COV-2 in oropharyngeal samples isolated from patients with suspected cases of COVID-19. The AGPC method of RNA extraction relies on simple chemical mixtures that can be easily obtained from commercial vendors or mixed by combining the needed chemicals. Overall the method is low-tech and routinely used in the many laboratories worldwide. The use for the AGPC method as a substitute for automated systems is especially important when automated systems are readily available but strained due to supply shortages. However, in places where these automated systems are not available, the AGPC method may allow for detection of viruses to levels comparable to those of the modern automated systems. This also means that a large number of scientists and technical personnel already are well-trained in using this method to address research questions for which RNA extraction is required. If necessary, these experienced personnel can be mobilized to aid in the RNA extraction, when either global demand reduces supply chains for critical components needed in the automated systems to extract RNA or if such automated systems are not readily available or overwhelmed in specific regions.

The AGPC method is robust and near the level of advanced commercial methods. When compared to the Maxwell^®^-based automation of RNA extraction using the same individual samples and the same SARS-COV-2 realtime PCR assay run in parallel, the results are virtually identical. For the Cobas^®^6800 system, the Ct values reported for the SARS-CoV-2 *E* gene were similarly comparable to those reported for the AGPC method. However, the Cobas^®^6800 system requires twice the volume of sample specimen for isolation of the RNA, utilizes approximately four times the amount of RNA product for the RT-qPCR analysis and is based upon proprietary primers and probes for detection of the SARS-CoV-2 *E* gene. Thus, a direct comparison cannot be made between the Cobas^®^6800 system and the results obtained with the used in-house SARS-COV-2 PCR assay. The detection sensitivity of the various methods is of utmost importance as viral titers may vary and false negative samples would allow individuals to spread COVID-19 infection inadvertently. A clear understanding of viral titers during the course of the infection is still lacking but may fluctuate during the course of the disease. While PCR based detection is not able to detect all, but nearly all COVID-19 cases (8, 9), it is important that the method used returns the highest amount of analytical accuracy in for diagnostic purposes. Our data suggest that the AGPC method is comparable in this respect.

While there are numerous advantages to the AGPC method, there are also inherent limitations. In comparison to automated RNA extraction systems there is extensive hands-on time and inadvertently risks of human errors. Furthermore, there is a possibility for loss of the sample, if the pellet is lost during the isolation step or if the sample is inadvertently mixed when transferring the aqueous phase between tubes. However, well established workflows can minimize these risks to very low levels. By spiking the sample specimens with the Nobilis ND C2 vaccine against Newcastle Disease containing inactivated virus, we obtained a good measure of the extraction efficiency and presence of potential inhibitors of the PCR analysis. This allowed us to monitor the entire extraction process and in the rare case of sample loss or other failures, repeat the RNA extraction and analysis, as only 200 μl out of the 1ml Eswab^TM^ media was used for the initial analysis.

RNA isolation using the AGPC method is favored among scientists for small scale RNA purification setups, due to its low cost, versatility and ease of use. Here we show that the AGPC method is easily scalable to volumes usable for clinical diagnostics as a supplement to conventional automated systems. The state-of-the-art Cobas^®^6800 system has a capacity of 384 samples per 8 hours, which is roughly equivalent to the throughput of our RNA isolation pipeline presented here. While the Cobas^®^6800 system is unsurpassed in ease, accuracy and sensitivity, the current COVID-19 pandemic and worldwide shortage of kits and reagents for automated systems, underlines the importance of redundant methods, which can be applied at a low cost, independent of proprietary reagents and commercial interest.

As with any work with viruses there is a chance of viral contamination. However, the collected sample specimens are mixed directly in organic solvent, which efficiently inactivates coronavirus. Hence, only this initial step needs to be carried out in a specialized class II facility (5, 10, 11). Nevertheless, AGPC solutions are hazardous and the isolation procedure in general needs to be carried out in fume hoods. This requires specialized workplaces commonly found across universities, hospitals, private research companies and institutions. Furthermore, if needed, many laboratories worldwide have RT-qPCR equipment that can be assembled at testing sites if necessary, for maintaining adequate PCR detection capacity.

Further experiments should aim to see whether the AGPC method could be optimized with respect to workflow, preparation time and to reduce manual handling of samples. Of relevance, would be to test whether swabs could be taken and directly placed in TRI Reagent^®^, if viral transport media is in short supply or if just to inactivate the virus and reduce the time spent aliquoting sample specimens into the organic solvent. Furthermore, similar comparisons could be made for other viruses to allow detection for various diagnostics in places not relying on automated systems.

## Data Availability

All data referred to in the manuscript will be available upon request to the corresponding author.

## Acknowledgements

We thank the laboratory and biomedical technicians at the Institute for Molecular Medicine, University of Southern Denmark and Department of Clinical Microbiology, Odense University Hospital for their expert technical assistance during testing, validation and implementation of the AGPC method to expand the SARS-CoV-2 testing capacity. We also thank dr. Kurt J. Handberg, Department of Clinical Microbiology, Skejby, Denmark, for kindly sharing primers and probe sequences for the detection of NDV.

## Ethical statement

The study described herein was conducted at the Department of Clinical Microbiology, Odense University Hospital, under the auspices of The Danish Health Authority. Exception from review by the ethical committee system and informed consent was given by the Regional Committees on Health Research Ethics for Southern Denmark in accordance with Danish law on assay development projects.

## Notes

### Competing Interest Statement

The authors have declared no competing interest.

### Clinical Trial

NA

### Funding Statement

No external funding was received

### Author Declarations

Exception from review by the Danish ethical committee system and informed consent was given by the Regional Committees on Health Research Ethics for Southern Denmark in accordance with Danish law on assay development projects.

